# Revealing associations between exposure to cyanobacteria toxins and COVID-19 outcomes in Colorado

**DOI:** 10.1101/2025.08.01.25332749

**Authors:** Anne E Thessen, Shawn T O’Neil, Melissa A Haendel, the National Clinical Cohort Collaborative (N3C) COVID Enclave

**Affiliations:** University of North Carolina Chapel Hill, Department of Genetics, Chapel Hill, North Carolina, USA

## Abstract

While environmental exposures are known to play a significant role in human disease, these effects are understudied compared to genomic and molecular components of disease. One example is the effect of chronic, low-level exposures to cyanobacteria toxins on health outcomes. Here we perform a retrospective analysis on real-world data in the National Clinical Cohort Collaborative (N3C) COVID Enclave, examining the possible impact of chronic exposure to cyanobacteria toxin on the severity of COVID-19 outcomes in patients from Colorado. We combined data from N3C, satellite data from the USEPA CyAN project, and field observations from the Colorado Department of Public Health & Environment (CDPHE). Our results show that COVID-19 patients living near recurring cyanobacteria blooms had 2.75 times higher odds of experiencing severe outcomes (hospitalization or death) than individuals who do not. In addition, living in a county with low to middle levels of poverty had protective effects. Further work is needed to understand the precise mechanism of action and fully understand the long-term risk of chronic exposures to low-level cyanobacteria toxins on health outcomes.

International Committee of Medical Journal Editors (ICMJE) Statement: Authorship was determined using ICMJE recommendations.

IRB: 24-2680

DUR ID: DUR-15EB88A

## Introduction

Many advances have been made in understanding genomic and molecular components of disease. The environmental component of disease is less well understood, but is known to play a significant role in cardiac, neurological, and immunological disease [1–3]. The effects of environmental algal toxins are not well understood, despite their impact on public health. In 2020, there were 227 harmful algal blooms reported in the US, resulting in 95 human and 1170 animal illnesses [4]. Most of these occurred in freshwater which had detectable amounts of microcystin, a toxin produced by a type of algae called cyanobacteria [4]. Toxin-producing cyanobacteria blooms are increasing globally and have caused many illnesses and deaths in humans and animals through an array of secondary metabolites that have hepatotoxic, neurotoxic, and skin-irritant properties [5,6]. Highly-visible acute mortality and illness events have been studied using field reports and laboratory investigations, but less is known about the public health consequences of repeated, low-level exposures to cyanobacterial toxins over long periods of time. This is partially due to their “silent” nature, but also due to the complexity of combining environmental and clinical data to investigate exposures of this type. Here, we combined clinical data from the National Clinical Cohort Collaborative (N3C) COVID Enclave [7] with water quality data from the USEPA CyAN [8] project to identify possible consequences of chronic cyanobacteria toxin exposure on COVID-19 outcomes.

The prevalence of toxin-producing cyanobacteria blooms in freshwater, including water used for drinking and recreation, has been increasing, caused by a combination of eutrophication and climate change [5,9,10]. The first documented deaths due to cyanobacteria toxins occurred in 1878 and involved cattle and sheep [11]. The first human intoxication event was recorded in 1931 when about 9,000 people developed acute gastroenteritis from cyanobacteria-contaminated drinking water [12]. Since that time, other reported symptoms in humans include skin rashes, skin and hepatic lesions, vomiting, headache, seizures, severe pneumonia, and death depending on the severity and mode of exposure. Several toxic events (resulting in death or illness) caused by cyanobacteria have been documented in multiple species, and it is well known that cyanobacteria produce hepatotoxins, neurotoxins, and skin irritants [6] (Table 1).

**Table 1.**
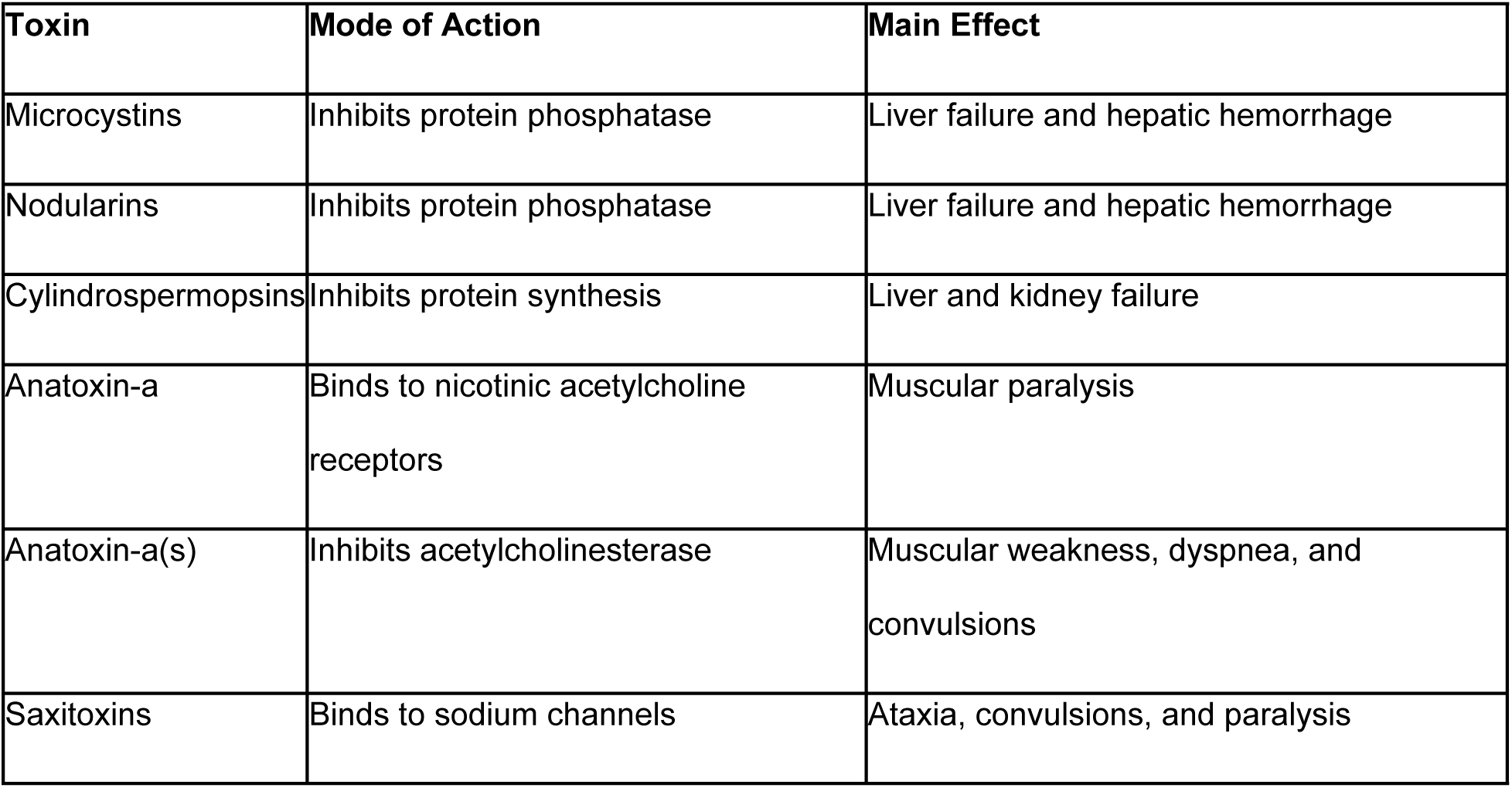
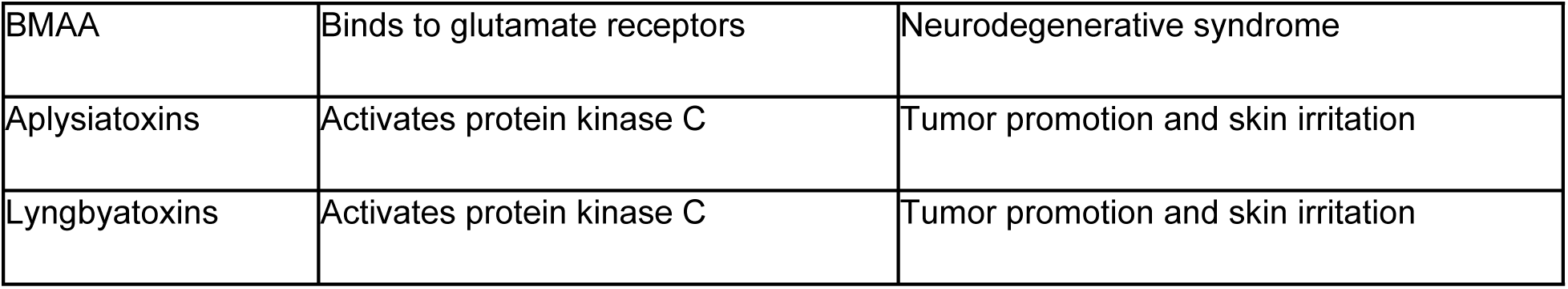
Cyanobacteria Toxins and Their Mode of Action. Recreated from [6].

| Toxin | Mode of Action | Main Effect |
| --- | --- | --- |
| Microcystins | Inhibits protein phosphatase | Liver failure and hepatic hemorrhage |
| Nodularins | Inhibits protein phosphatase | Liver failure and hepatic hemorrhage |
| Cylindrospermopsins | Inhibits protein synthesis | Liver and kidney failure |
| Anatoxin-a | Binds to nicotinic acetylcholine receptors | Muscular paralysis |
| Anatoxin-a(s) | Inhibits acetylcholinesterase | Muscular weakness, dyspnea, and convulsions |
| Saxitoxins | Binds to sodium channels | Ataxia, convulsions, and paralysis |
| BMAA | Binds to glutamate receptors | Neurodegenerative syndrome |
| Aplysiatoxins | Activates protein kinase C | Tumor promotion and skin irritation |
| Lyngbyatoxins | Activates protein kinase C | Tumor promotion and skin irritation |

Cyanobacteria toxins can be inhaled as an aerosol [13,14] and ingested in water or food [10]. The presence of cyanobacteria and their toxins in bioaerosols has been well documented [15,16], and exposure to toxin-containing aerosols may not be restricted to areas near an affected water body [17]. A previous study in California suggested that while proximity to a cyanobacteria bloom resulted in inhaling aerosolized cyanobacteria toxins, it was unlikely to cause adverse acute effects in healthy individuals over the short term [13]. Laboratory studies demonstrated that cyanobacteria aerosols can cause irritation and inflammatory injuries in human upper respiratory airway epithelial cells [18]. A study examining cyanobacteria in the human respiratory tract found no relationship between the presence of cyanobacteria in a patient and that patient’s proximity to a water body [17]. Field data (*in situ* samplers) suggest that cyanobacteria toxins can travel over a mile from source [19] and can be found in indoor environments [15]. Controlled studies show that cyanobacteria toxins quickly decay in sunlight [20] and models suggest that cyanobacteria toxins are only able to travel approximately half a mile from source before decaying [21].

Less understood is the relationship between chronic disease and repeated exposure to cyanobacteria toxins at levels too low to cause acute illness. Repeated exposure to low-levels of cyanobacteria toxins may occur via food, absorption through the skin, or inhalation. Several epidemiological studies have found a link between the presence of cyanobacteria toxins in drinking water and liver cancer around the world [22–25]. Increasing evidence suggests that environmental toxins play an important role in liver cancer development and could be contributing to the increase in chronic liver disease [26]. Several studies suggest that aerosolized cyanobacteria toxins can be inhaled [27] and it has been hypothesized that exposure through inhalation or ingestion is connected to neurodegenerative disease [28–31]. In addition to being directly harmful, the tissue damage caused by cyanobacteria toxins could potentially increase susceptibility to other diseases, including COVID-19. Studies have shown that patients with chronic respiratory, kidney, liver, and neurological disease were at higher risk of hospitalization due to COVID-19 [27]. Exact amounts of toxin that result in organ damage, and lag time between exposure and organ damage without acute illness in humans is unknown.

Climate change, in combination with eutrophication and land use change is expected to exacerbate the duration, frequency, and size of cyanobacteria blooms [5,32] partially because of the anticipated increases in temperature and drought (leading to more stagnant water bodies). A report describing the specific impacts of climate change in Colorado predicted (with high confidence) increases in temperature and more frequent and intense droughts, but impacts involving cyanobacteria were not discussed [33]. Toxin-producing cyanobacteria are already a common occurrence in Colorado water bodies during warm months and these predicted climate trends suggest that blooms will become more frequent in the future. Recent work in Europe showed that climate change impacts cool lakes (high latitude) and warm lakes (low latitude) differently and found that cyanobacteria blooms in the high latitude lakes produced more toxin [34]. In some contexts, ecosystems in high latitude and high altitude environments share similar characteristics e.g., cold temperatures and short growing seasons, but It is not clear if results from high latitude European lakes can be applied to high altitude Colorado lakes.

There are approximately 1,533 public lakes in Colorado with over 10 surface acres in size [35]. Cyanobacteria naturally occur in Colorado waterways and only become a public health threat when they produce toxins. Only some of the water bodies in Colorado are routinely monitored for the presence of cyanobacteria and their toxins. These data, from the Colorado Department of Public Health & Environment (CDPHE) from 2014 to present, show that toxin-producing cyanobacteria blooms have been documented across a wide swath of the state. The CyAN project provides satellite-derived cyanobacteria concentrations for water bodies in the continental United States, but this project has only been available since 2020 and is limited to larger water bodies [25,26]. Cyanobacteria blooms have been documented in Colorado reservoirs [36,37] and in the artificial Sheldon Lake [38]. Earlier in 2022, the Pikeview Reservoir was removed as a municipal water source due to cyanobacteria [39]. While municipal water treatment plants have some strategies to remove cyanobacteria and their toxins, home water filtration units typically are not able to [40]. In 2023 and 2024, several cyanobacteria blooms were documented in multiple water bodies in Colorado, some of which resulted in closure to recreational use [41–43]. The exact history of the prevalence of toxic cyanobacteria in Colorado waterways is not well known due to a lack of long-term, routine monitoring. Recently, however, advances in satellite monitoring combined with *in situ* measurements make an investigation into public health effects plausible.

## Materials & Methods

### Identification of High Risk Water Bodies and Zip Codes

We requested data from the CyAN project by manually searching for and choosing water bodies in Colorado using the CyAN web application and making a data request [44]. CyAN data were accessed 25 May 2023. Data from the CDPHE were gathered from their website the same month [45]. Combining these two partially overlapping data sources gave information for 276 water bodies (Fig. 1; <u>Supp Data</u> 1). A water body was identified as having chronic cyanobacteria blooms if: cyanobacteria cell concentrations reached 10^6^ cells per mL at least once, cyanobacteria cells were observed for at least three years, and, if tested, toxins were found. Seventy-seven water bodies met these criteria. Maps of zip codes and aerial imagery were used to identify the zip codes that had at least 95% of its residential population within ∼5 km of these 77 water bodies (Fig. 1). These were the “high risk” zip codes. “Low risk” zip codes were identified as having the vast majority of its residential population either not near any water body or only near moving water (<u>Supp Data 2</u>).

**Figure 1:**
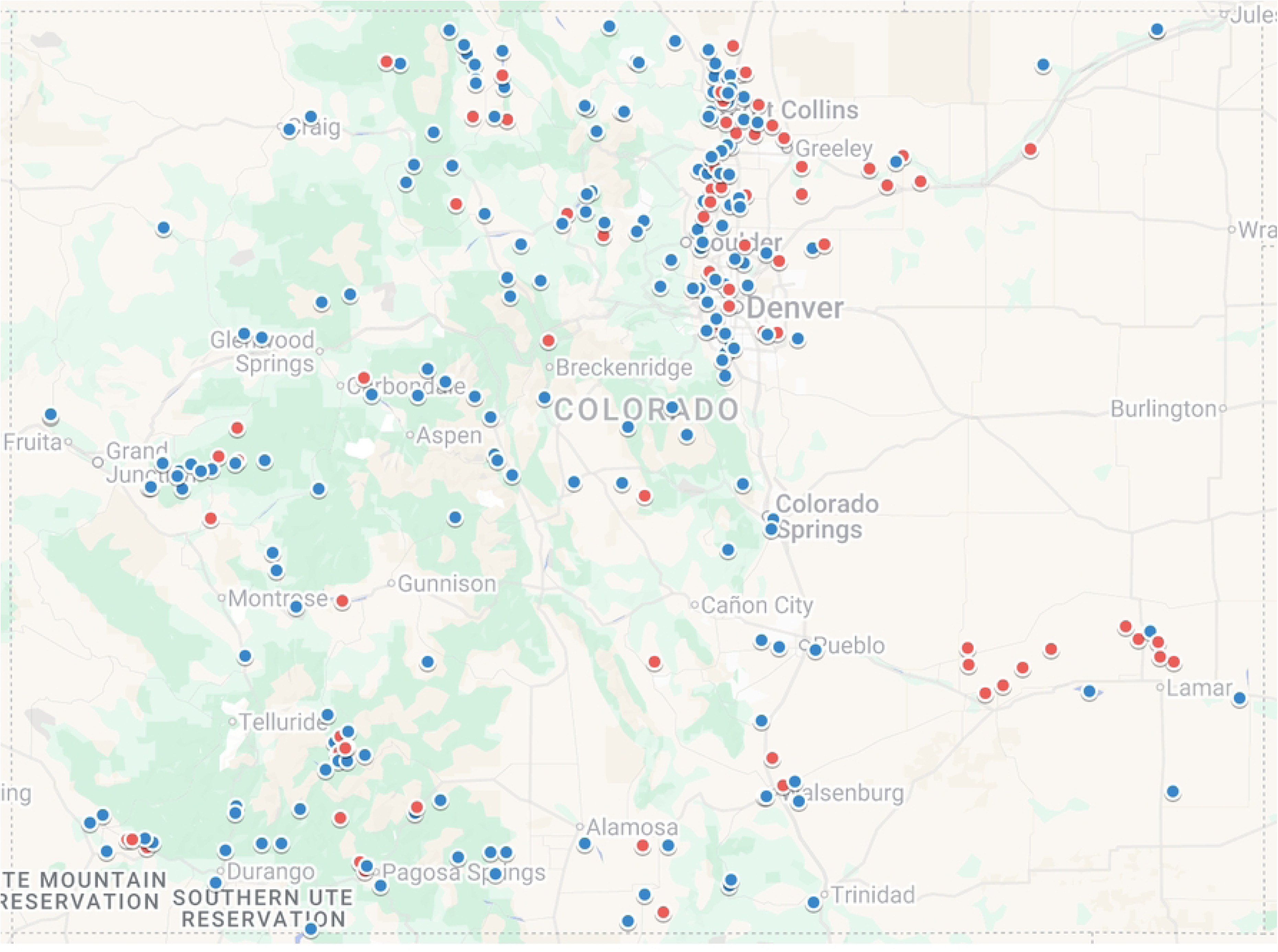
Map of Colorado Water Bodies Examined in this Study. Water bodies were labeled as being low risk for cyanobacteria toxin exposure (blue) or high risk for cyanobacteria toxin exposure (red) based on data from CyAN and CDPHE. Three low-risk water bodies are not on the map because we were unable to get accurate coordinates. Figure made using Google MyMaps.

### Identification of COVID Patients

The N3C Enclave is a secure platform through which harmonized clinical data provided by contributing members of N3C are stored [7]. The Enclave includes demographic and clinical characteristics of patients who have been tested for or diagnosed with COVID-19, and further information about the strategies and outcomes of treatments for those suspected or confirmed to have the virus. The N3C Enclave is available for public research use. To access data including that used in this manuscript, institutions must have a signed Data Use Agreement executed with the U.S. National Center for Advancing Translational Sciences (NCATS) and their investigators must complete mandatory training and submit a Data Use Request (DUR) to N3C. To request N3C data access, researchers must follow instructions at https://covid.cd2h.org/onboarding. This project can be replicated within the N3C Enclave by fully onboarded, trained users with an approved DUR. More than 4,000 researchers currently have access to data in N3C; together they represent more than 300 US research institutions. Code used in this analysis and all concept sets are available at [46]. Reviewers can confidentially request access by these same means.

The N3C data transfer to NCATS is performed under a Johns Hopkins University Reliance Protocol # IRB00249128 or individual site agreements with NIH. The work presented here was performed under a University of Colorado Anschutz Medical Campus (23-1860) and University of North Carolina Chapel Hill (24-2680) IRB protocols. A waiver of informed consent was approved by the NIH Intramural IRB for storing, operational management, and maintaining data within the N3C Enclave. This was done under the HHS regulations at 45 CFR 46. The need for consent was waived by the ethics committee. This study did not include minors.

N3C hosts a very large collection of de-identified electronic health records in the OMOP Common Data Model (CDM), with data available beginning 1/1/2018. Analyses were performed on N3C Level 3 data from release v185 (10/10/2024) representing over 23.3 million patients from 85 pseudo-anonymous data contributing sites across the United States [7,47]. This data set contained two HIPAA identifiers; thus, some individuals might be identifiable after data collection with extra work, which was forbidden by the data use agreement. We filtered to include only patients associated with a defined high-risk or low-risk zip code, effectively restricting to 45 data contributors. We considered only COVID-19 positive patients: those with a positive PCR test, Antigen test, or diagnosis. The first instance of these was used to identify primary COVID-19 infection date and subsequent severity as either hospitalization, Emergency Department presentation (with accompanying positive test or diagnosis), or death within 60 days of primary diagnosis. Data were filtered to include only patients associated with defined high-risk (1) or low-risk zip codes (0), and also excluded smokers, pregnant patients, minors, and those with diabetes, obesity, heart disease, organ transplant, or stroke prior to primary COVID-19 infection (Fig. 2).

**Figure 2:**
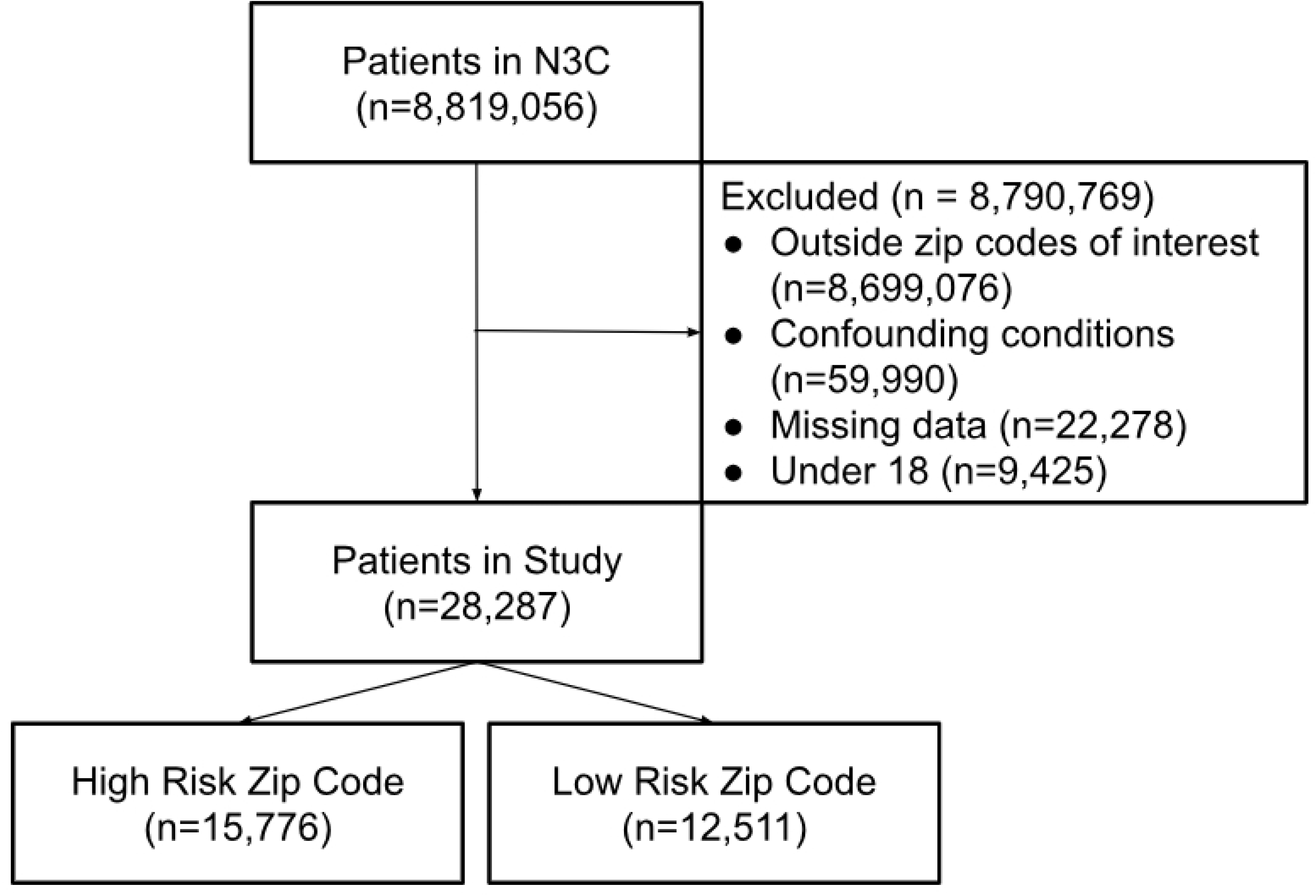
Consort Flow Diagram. This study started with over 8 million patients in N3C and Identified approximately 28K patients that met the study criteria.

Patient covariates included vaccination status (1 or more prior to primary infection), discretized age (adult 19-65, senior >65), discretized county-level poverty rate from 2019 American Community Survey data [48] (low 0-5%, middle 5-15%, high >15%), and discretized healthcare access from the 2020 Sharecare-BUSPH Social Determinants of Health Index [49] (low 0-1.5 MDs per thousand population, reasonable 1.5-2.5, strong >2.5). To allow sufficient group sizes for analysis, race and ethnicity were included and combined into categories Latino (Hispanic or Latino ethnicity), White (race White), Black (race Black or African American), and Other (race Asian, American Indian or Alaska Native, or Native Hawaiian or Other Pacific Islander.) Patients without complete covariate data were excluded from further analyses. Supplemental table 3 describes patient group counts, ranging from <20 individuals to 5,876 individuals.

### Statistical Analysis

All statistical analyses were performed in R v4.4.2. High- and low-risk patients were matched with ATT as the estimand, using exact matching on age category, race, and sex using MatchIt v4.6.0 [50]. Binary COVID-19 severity was modeled with generalized linear logistic regression, weighted with with stratum propensity score weights using glm() in R, using formula severity_type ∼ cyano_risk * (vaccine_status + poverty_category + access_category). Odds Ratios (ORs) were computed using reported model coefficients.

## Results

This study was performed using data from ∼28k patients and was well balanced across sex, age, and high vs low cyanobacteria toxin risk (Table 2). There were many more patients who did not have a severe COVID-19 outcome than did.

**Table 2:** Patient Demographics.

|  |  |
| --- | --- |
| Number | 28,287 |
| Male / Female | 12,236 / 16,051 |
| Age | 45.8 +/- 17.9 |
| Severe COVID / Not severe COVID | 3,570 / 24,717 |
| High Risk / Low Risk | 15,776 / 12,511 |

Compared to the baseline group (low risk, unvaccinated, high poverty, low care access), patients in high risk areas were significantly more likely to experience a severe COVID-19 outcome (OR 2.7, p<0.0001) (<u>Supp Data 4</u>, Fig 3). Model coefficients also suggest that vaccinated patients were less likely to have a severe COVID outcome, which is supported by prior knowledge [51,52]. There also appear to be interactions that reduce COVID severity for wealthy and middle-class patients living in high cyanobacteria risk areas [52,53].

**Figure 3:**
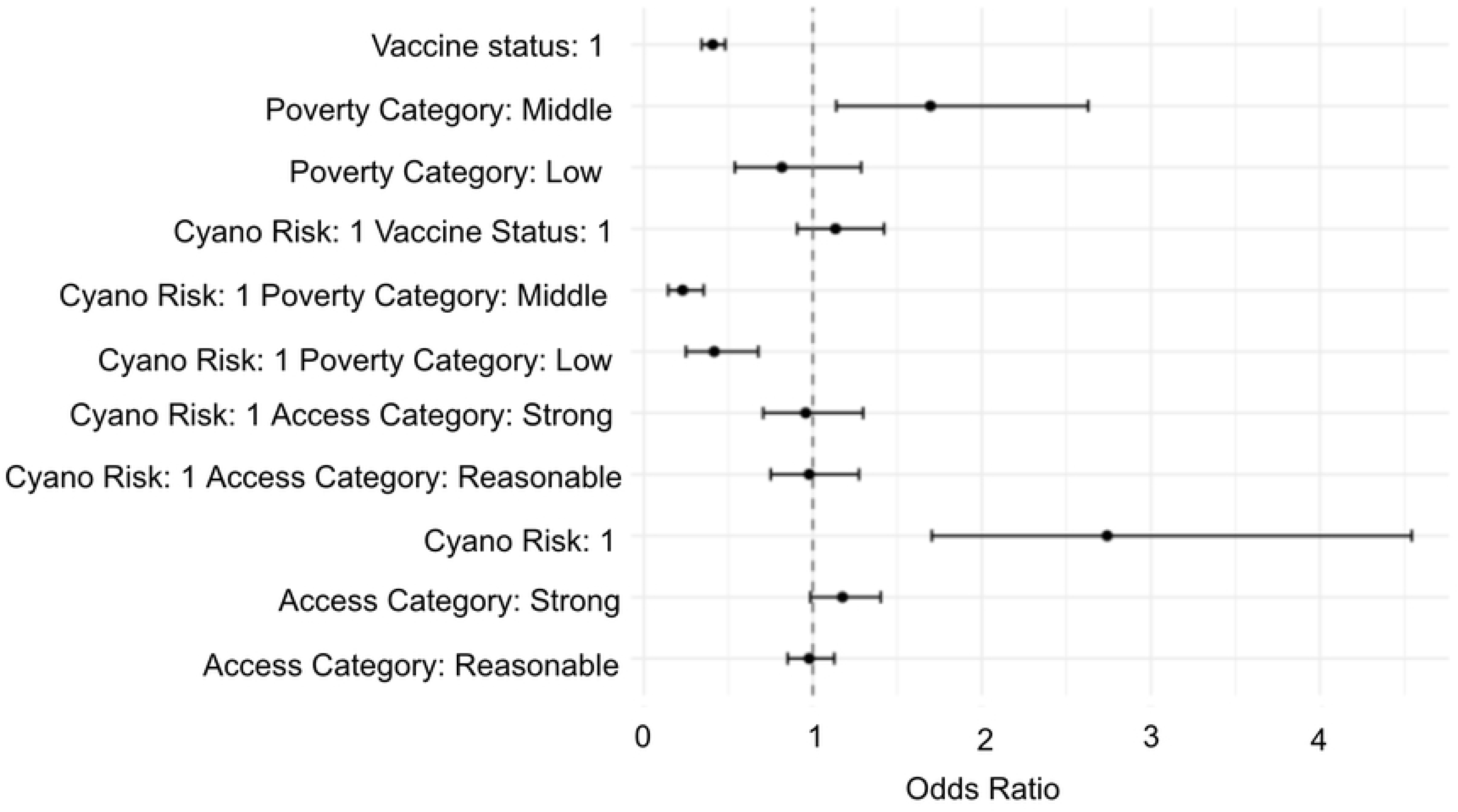
Forest Plot of Odds Ratio from GLM. Patients living in zip codes identified as high risk for chronic exposure to low levels of cyanobacteria toxin were more likely to have a severe COVID-19 outcome. There are interactions that reduce COVID-19 severity for patients living in high risk zip codes that are also wealthy or middle-class.

## Discussion

The World Health Organization estimated that 3.4 million people died during the COVID-19 pandemic [54] and, as of the time of writing, thousands per month continue to die, mostly in the USA [55]. Factors that impact COVID severity thus remain impactful areas of research. The Centers for Disease Control identified several underlying medical conditions (e.g., obesity and chronic kidney disease) and demographic factors (e.g., age and race) that increase the likelihood of a severe COVID outcome [56]. Additionally, several ACE2 receptor gene variants in humans have been identified as impacting COVID susceptibility, and prevalence of these variants differs by ethnic group [57–59]. There is indirect evidence of some environmental exposures increasing the likelihood of a severe COVID outcome, such as air pollution (particulate matter, NO2, ozone), heavy metals, and endocrine disruptors [60]. Direct evidence of the impact and specific mechanism of action of a chemical or pollutant on COVID severity has not been firmly established.

Chronic exposure to low-levels of aerosolized cyanobacteria toxins could increase likelihood of a severe COVID outcome by impairing respiratory function. It is well established that comorbidities impairing respiratory function, such as asthma and chronic lung disease, can make a patient more likely to have a severe COVID-19 outcome [61–63]. While our results suggest this mechanism of action, this study did not look for direct evidence of impaired lung function in areas of high risk of exposure to cyanobacteria toxins. The complete effects of spending significant amounts of time near water bodies with chronic cyanobacteria blooms is not yet clear.

This work examines an understudied environmental exposure, chronic low-level exposure to cyanobacteria toxins, and its impact on public health. These findings suggest that in addition to the direct impact of the toxins on organ and tissue function, chronic, low-levels of cyanobacteria toxins can increase an individual’s susceptibility to some communicable disease. Further study is needed to see if these results are applicable to a larger population and to identify the specific mechanism by which proximity to recurring cyanobacteria blooms leads to more severe COVID-19 outcomes.

## Data Availability

The N3C Enclave is available for public research use. To access data including that used in this manuscript, institutions must have a signed Data Use Agreement executed with the U.S. National Center for Advancing Translational Sciences (NCATS) and their investigators must complete mandatory training and submit a Data Use Request (DUR) to N3C. To request N3C data access, researchers must follow instructions at https://covid.cd2h.org/onboarding. This project can be replicated within the N3C Enclave by fully onboarded, trained users with an approved DUR. Reviewers can confidentially request access by these same means.

## Acknowledgements

The analyses described in this publication were conducted with data and tools accessed through the NCATS N3C Data Enclave https://covid.clinicalcohort.org and N3C Attribution & Publication Policy v 1.2-2020-08-25b created through the Center for Data to Health. This research was possible because of the patients whose information is included within the data and the organizations (https://ncats.nih.gov/n3c/resources/data-contribution/data-transfer-agreement-signatories) and scientists who have contributed to the on-going development of this community resource [7].

Disclaimer The N3C Publication committee confirmed that this manuscript msid:2464.857 is in accordance with N3C data use and attribution policies; however, this content is solely the responsibility of the authors and does not necessarily represent the official views of the National Institutes of Health or the N3C program.

## IRB

The N3C data transfer to NCATS is performed under a Johns Hopkins University Reliance Protocol # IRB00249128 or individual site agreements with NIH. The N3C Data Enclave is managed under the authority of the NIH; information can be found at https://ncats.nih.gov/n3c/resources.

We gratefully acknowledge the following core contributors to N3C: Adam B. Wilcox, Adam M. Lee, Alexis Graves, Alfred (Jerrod) Anzalone, Amin Manna, Amit Saha, Amy Olex, Andrea Zhou, Andrew E. Williams, Andrew M. Southerland, Andrew T. Girvin, Anita Walden, Anjali Sharathkumar, Benjamin Amor, Benjamin Bates, Brian Hendricks, Brijesh Patel, G. Caleb Alexander, Carolyn T. Bramante, Cavin Ward-Caviness, Charisse Madlock-Brown, Christine Suver, Christopher G. Chute, Christopher Dillon, Chunlei Wu, Clare Schmitt, Cliff Takemoto, Dan Housman, Davera Gabriel, David A. Eichmann, Diego Mazzotti, Donald E. Brown, Eilis Boudreau, Elaine L. Hill, Emily Carlson Marti, Emily R. Pfaff, Evan French, Farrukh M Koraishy, Federico Mariona, Fred Prior, George Sokos, Greg Martin, Harold P. Lehmann, Heidi Spratt, Hemalkumar B. Mehta, J.W. Awori Hayanga, Jami Pincavitch, Jaylyn Clark, Jeremy Richard Harper, Jessica Yasmine Islam, Jin Ge, Joel Gagnier, Johanna J. Loomba, John B. Buse, Jomol Mathew, Joni L. Rutter, Julie A. McMurry, Justin Guinney, Justin Starren, Karen Crowley, Katie Rebecca Bradwell, Kellie M. Walters, Ken Wilkins, Kenneth R. Gersing, Kenrick Cato, Kimberly Murray, Kristin Kostka, Lavance Northington, Lee Pyles, Lesley Cottrell, Lili M. Portilla, Mariam Deacy, Mark M. Bissell, Marshall Clark, Mary Emmett, Matvey B. Palchuk, Melissa A. Haendel, Meredith Adams, Meredith Temple-O’Connor, Michael G. Kurilla, Michele Morris, Nasia Safdar, Nicole Garbarini, Noha Sharafeldin, Ofer Sadan, Patricia A. Francis, Penny Wung Burgoon, Philip R.O. Payne, Randeep Jawa, Rebecca Erwin-Cohen, Rena C. Patel, Richard A. Moffitt, Richard L. Zhu, Rishikesan Kamaleswaran, Robert Hurley, Robert T. Miller, Saiju Pyarajan, Sam G. Michael, Samuel Bozzette, Sandeep K. Mallipattu, Satyanarayana Vedula, Scott Chapman, Shawn T. O’Neil, Soko Setoguchi, Stephanie S. Hong, Steven G. Johnson, Tellen D. Bennett, Tiffany J. Callahan, Umit Topaloglu, Valery Gordon, Vignesh Subbian, Warren A. Kibbe, Wenndy Hernandez, Will Beasley, Will Cooper, William Hillegass, Xiaohan Tanner Zhang. Details of contributions available at covid.cd2h.org/core-contributors

## Data Partners with Released Data

The following institutions whose data is released or pending: Available: Advocate Health Care Network — UL1TR002389: The Institute for Translational Medicine (ITM) • Aurora Health Care Inc — UL1TR002373: Wisconsin Network For Health Research • Boston University Medical Campus — UL1TR001430: Boston University Clinical and Translational Science Institute • Brown University — U54GM115677: Advance Clinical Translational Research (Advance-CTR) • Carilion Clinic — UL1TR003015: iTHRIV Integrated Translational health Research Institute of Virginia • Case Western Reserve University — UL1TR002548: The Clinical & Translational Science Collaborative of Cleveland (CTSC) • Charleston Area Medical Center — U54GM104942: West Virginia Clinical and Translational Science Institute (WVCTSI) • Children’s Hospital Colorado — UL1TR002535: Colorado Clinical and Translational Sciences Institute • Columbia University Irving Medical Center — UL1TR001873: Irving Institute for Clinical and Translational Research • Dartmouth College — None (Voluntary) Duke University — UL1TR002553: Duke Clinical and Translational Science Institute • George Washington Children’s Research Institute — UL1TR001876: Clinical and Translational Science Institute at Children’s National (CTSA-CN) • George Washington University — UL1TR001876: Clinical and Translational Science Institute at Children’s National (CTSA-CN) • Harvard Medical School — UL1TR002541: Harvard Catalyst • Indiana University School of Medicine — UL1TR002529: Indiana Clinical and Translational Science Institute • Johns Hopkins University — UL1TR003098: Johns Hopkins Institute for Clinical and Translational Research • Louisiana Public Health Institute — None (Voluntary) • Loyola Medicine — Loyola University Medical Center • Loyola University Medical Center — UL1TR002389: The Institute for Translational Medicine (ITM) • Maine Medical Center — U54GM115516: Northern New England Clinical & Translational Research (NNE-CTR) Network • Mary Hitchcock Memorial Hospital & Dartmouth Hitchcock Clinic — None (Voluntary) • Massachusetts General Brigham — UL1TR002541: Harvard Catalyst • Mayo Clinic Rochester — UL1TR002377: Mayo Clinic Center for Clinical and Translational Science (CCaTS) • Medical University of South Carolina — UL1TR001450: South Carolina Clinical & Translational Research Institute (SCTR) • MITRE Corporation — None (Voluntary) • Montefiore Medical Center — UL1TR002556: Institute for Clinical and Translational Research at Einstein and Montefiore • Nemours — U54GM104941: Delaware CTR ACCEL Program • NorthShore University HealthSystem — UL1TR002389: The Institute for Translational Medicine (ITM) • Northwestern University at Chicago — UL1TR001422: Northwestern University Clinical and Translational Science Institute (NUCATS) • OCHIN — INV-018455: Bill and Melinda Gates Foundation grant to Sage Bionetworks • Oregon Health & Science University — UL1TR002369: Oregon Clinical and Translational Research Institute • Penn State Health Milton S. Hershey Medical Center — UL1TR002014: Penn State Clinical and Translational Science Institute • Rush University Medical Center — UL1TR002389: The Institute for Translational Medicine (ITM) • Rutgers, The State University of New Jersey — UL1TR003017: New Jersey Alliance for Clinical and Translational Science • Stony Brook University — U24TR002306 • The Alliance at the University of Puerto Rico, Medical Sciences Campus — U54GM133807: Hispanic Alliance for Clinical and Translational Research (The Alliance) • The Ohio State University — UL1TR002733: Center for Clinical and Translational Science • The State University of New York at Buffalo — UL1TR001412: Clinical and Translational Science Institute • The University of Chicago — UL1TR002389: The Institute for Translational Medicine (ITM) • The University of Iowa — UL1TR002537: Institute for Clinical and Translational Science • The University of Miami Leonard M. Miller School of Medicine — UL1TR002736: University of Miami Clinical and Translational Science Institute • The University of Michigan at Ann Arbor — UL1TR002240: Michigan Institute for Clinical and Health Research • The University of Texas Health Science Center at Houston — UL1TR003167: Center for Clinical and Translational Sciences (CCTS) • The University of Texas Medical Branch at Galveston — UL1TR001439: The Institute for Translational Sciences • The University of Utah — UL1TR002538: Uhealth Center for Clinical and Translational Science • Tufts Medical Center — UL1TR002544: Tufts Clinical and Translational Science Institute • Tulane University — UL1TR003096: Center for Clinical and Translational Science • The Queens Medical Center — None (Voluntary) • University Medical Center New Orleans — U54GM104940: Louisiana Clinical and Translational Science (LA CaTS) Center • University of Alabama at Birmingham — UL1TR003096: Center for Clinical and Translational Science • University of Arkansas for Medical Sciences — UL1TR003107: UAMS Translational Research Institute • University of Cincinnati — UL1TR001425: Center for Clinical and Translational Science and Training • University of Colorado Denver, Anschutz Medical Campus — UL1TR002535: Colorado Clinical and Translational Sciences Institute • University of Illinois at Chicago — UL1TR002003: UIC Center for Clinical and Translational Science • University of Kansas Medical Center — UL1TR002366: Frontiers: University of Kansas Clinical and Translational Science Institute • University of Kentucky — UL1TR001998: UK Center for Clinical and Translational Science • University of Massachusetts Medical School Worcester — UL1TR001453: The UMass Center for Clinical and Translational Science (UMCCTS) • University Medical Center of Southern Nevada — None (voluntary) • University of Minnesota — UL1TR002494: Clinical and Translational Science Institute • University of Mississippi Medical Center — U54GM115428: Mississippi Center for Clinical and Translational Research (CCTR) • University of Nebraska Medical Center — U54GM115458: Great Plains IDeA-Clinical & Translational Research • University of North Carolina at Chapel Hill — UL1TR002489: North Carolina Translational and Clinical Science Institute • University of Oklahoma Health Sciences Center — U54GM104938: Oklahoma Clinical and Translational Science Institute (OCTSI) • University of Pittsburgh — UL1TR001857: The Clinical and Translational Science Institute (CTSI) • University of Pennsylvania — UL1TR001878: Institute for Translational Medicine and Therapeutics • University of Rochester — UL1TR002001: UR Clinical & Translational Science Institute • University of Southern California — UL1TR001855: The Southern California Clinical and Translational Science Institute (SC CTSI) • University of Vermont — U54GM115516: Northern New England Clinical & Translational Research (NNE-CTR) Network • University of Virginia — UL1TR003015: iTHRIV Integrated Translational health Research Institute of Virginia • University of Washington — UL1TR002319: Institute of Translational Health Sciences • University of Wisconsin-Madison — UL1TR002373: UW Institute for Clinical and Translational Research • Vanderbilt University Medical Center — UL1TR002243: Vanderbilt Institute for Clinical and Translational Research • Virginia Commonwealth University — UL1TR002649: C. Kenneth and Dianne Wright Center for Clinical and Translational Research • Wake Forest University Health Sciences — UL1TR001420: Wake Forest Clinical and Translational Science Institute • Washington University in St. Louis — UL1TR002345: Institute of Clinical and Translational Sciences • Weill Medical College of Cornell University — UL1TR002384: Weill Cornell Medicine Clinical and Translational Science Center • West Virginia University — U54GM104942: West Virginia Clinical and Translational Science Institute (WVCTSI) Submitted: Icahn School of Medicine at Mount Sinai — UL1TR001433: ConduITS Institute for Translational Sciences • The University of Texas Health Science Center at Tyler — UL1TR003167: Center for Clinical and Translational Sciences (CCTS) • University of California, Davis — UL1TR001860: UCDavis Health Clinical and Translational Science Center • University of California, Irvine — UL1TR001414: The UC Irvine Institute for Clinical and Translational Science (ICTS) • University of California, Los Angeles — UL1TR001881: UCLA Clinical Translational Science Institute • University of California, San Diego — UL1TR001442: Altman Clinical and Translational Research Institute • University of California, San Francisco — UL1TR001872: UCSF Clinical and Translational Science Institute NYU Langone Health Clinical Science Core, Data Resource Core, and PASC Biorepository Core — OTA-21-015A: Post-Acute Sequelae of SARS-CoV-2 Infection Initiative (RECOVER) Pending: Arkansas Children’s Hospital — UL1TR003107: UAMS Translational Research Institute • Baylor College of Medicine — None (Voluntary) • Children’s Hospital of Philadelphia — UL1TR001878: Institute for Translational Medicine and Therapeutics • Cincinnati Children’s Hospital Medical Center — UL1TR001425: Center for Clinical and Translational Science and Training • Emory University — UL1TR002378: Georgia Clinical and Translational Science Alliance • HonorHealth — None (Voluntary) • Loyola University Chicago — UL1TR002389: The Institute for Translational Medicine (ITM) • Medical College of Wisconsin — UL1TR001436: Clinical and Translational Science Institute of Southeast Wisconsin • MedStar Health Research Institute — None (Voluntary) • Georgetown University — UL1TR001409: The Georgetown-Howard Universities Center for Clinical and Translational Science (GHUCCTS) • MetroHealth — None (Voluntary) • Montana State University — U54GM115371: American Indian/Alaska Native CTR • NYU Langone Medical Center — UL1TR001445: Langone Health’s Clinical and Translational Science Institute • Ochsner Medical Center — U54GM104940: Louisiana Clinical and Translational Science (LA CaTS) Center • Regenstrief Institute — UL1TR002529: Indiana Clinical and Translational Science Institute • Sanford Research — None (Voluntary) • Stanford University — UL1TR003142: Spectrum: The Stanford Center for Clinical and Translational Research and Education • The Rockefeller University — UL1TR001866: Center for Clinical and Translational Science • The Scripps Research Institute — UL1TR002550: Scripps Research Translational Institute • University of Florida — UL1TR001427: UF Clinical and Translational Science Institute • University of New Mexico Health Sciences Center — UL1TR001449: University of New Mexico Clinical and Translational Science Center • University of Texas Health Science Center at San Antonio — UL1TR002645: Institute for Integration of Medicine and Science • Yale New Haven Hospital — UL1TR001863: Yale Center for Clinical Investigation

## Supporting Information Captions

S1 Data: Colorado Waterbody and Cyanobacteria Data Summary

S2 Data: High Risk and Low Risk Zip Codes Used in This Study

S3 Data: Patient Demographic Summary

S4 Data: GLM Results and Odds Ratios

